# Refined fully automated RT-qPCR assay for simultaneous detection of SARS-CoV-2, Influenza A/B, and RSV with target optimization for improved variant resilience

**DOI:** 10.1101/2025.10.29.25339068

**Authors:** Susanne Pfefferle, Dominik Nörz, Katja Giersch, Moritz Grunwald, Hui Ting Tang, Lisa Sophie Pflüger, Marc Lütgehetmann

**Affiliations:** Institute of Medical Microbiology, Virology and Hygiene, University Medical Centre Hamburg-Eppendorf (UKE), Hamburg, Germany

**Keywords:** Respiratory infection, real time polymerase chain reaction, high-throughput automated detection, LDT, cobas 5800, cobas 6800, cobas 8800, molecular diagnostics, SARS-CoV-2, influenza, RSV

## Abstract

**Background:** Continuous viral evolution is likely to cause a loss of performance or failure of existing diagnostic assays. Here, we delineate the adaptation and validation of an operational laboratory developed (LDT) multiplex RT-qPCR for simultaneous detection of SARS-CoV-2, influenza A/B (FluA/B) and respiratory syncytial virus (RSV) on a high-throughput, fully automated platform. Furthermore, we explore the integration of an alternative SARS-CoV-2 target to enhance assay robustness

**Methods:** An adaption of the operational assay (PMID:38820916) was performed and a novel SARS-CoV-2 target (macrodomain, Mac1) was implemented. Analytical performance of the adapted assay was evaluated using digital-PCR based standards or international reference material and clinical performance was assessed on clinical samples with a CE-IVD comparator assay.

**Results:** Analytical sensitivity (lower limit of detection (LoD)) was 70.9 IU/ml for SARS-CoV-2, and 112-474, 919 and 1,720 cp/ml for influenza A, influenza B and RSV, respectively, with linear ranges of 26.3-36.4 ct (SARS-CoV-2), 26.8-37.7 ct (FluA), 28.5-37.9 (FluB) and 25.6-38.2 (RSV). Clinical performance evaluation confirmed improved performance (e.g. FluA/B detection -1.6/-4.81 ct) and comparable performance to the CE-IVD assay (excellent correlation of the SARS-CoV-2 assays, more effective detection of influenza B). Overall, positive/negative agreement was 94%/96% (SARS-CoV-2), and 100%/100% (FluA/B, RSV).

**Conclusion:** The adapted LDT assay as a focused syndromic assay provides reliable detection of major respiratory viruses on a high-throughput platform. The strategic targeting of conserved genomic regions ensures diagnostic resilience, while automated integration facilitates scalable laboratory operations. This approach facilitates robust pathogen surveillance and expedites clinical decision-making during periods of co-circulation and epidemic surge.

## Introduction

The COVID-19 pandemic underscored the need for rapid, scalable molecular diagnostics for respiratory viruses [1,2]. RT-qPCR remains the gold standard, and automated high-throughput platforms enable laboratories to meet clinical and public health demands. Concurrently, the major respiratory viral pathogens SARS-CoV-2, influenza A/B and RSV continue to burden healthcare systems [3–5] making robust multiplex diagnostics essential, particularly during co-circulation periods.

Genetic variability poses a major diagnostic challenge, as viral sequence diversity can introduce mutations in primer-probe binding sites, reducing assay sensitivity [6]. Targeting conserved, functionally constrained genomic regions mitigates evolutionary escape.

Genetic variability poses a major diagnostic challenge, as viral sequence diversity can introduce mutations in primer-probe binding sites, reducing assay performance [6]. Targeting conserved, functionally constrained genomic regions mitigates evolutionary escape.

We previously reported a multiplexed RT-qPCR assay for SARS-CoV-2, influenza A/B, and RSV on the Cobas platform [7]. During routine evaluation of the operational assay we observed slight performance losses, prompting assay redesign. We identified the SARS-CoV-2 Mac1 domain as a promising new target due to its conservation and critical role in viral replication [8–10].

Here, we describe the updated fully automated multiplexed RT-qPCR assay with redesigned primer-probe sets. We validated analytical sensitivity and inclusivity against contemporary viral lineages and assessed clinical performance in routine high-throughput operations. This updated assay provides enhanced variant resilience while maintaining seamless integration into existing laboratory workflows.

## Methods

### Assay redesign

For the new assay (hereafter RESP1_V1), primers and probes were adapted or exchanged for optimal inclusivity, reduced oligo interaction and system compatibility as described [7] (Table 1). In-silico inclusivity was evaluated using GenBank queries and Geneious Prime alignments (version 2025.1.2). Oligonucleotide details are in Table 2.

**Table 1.**
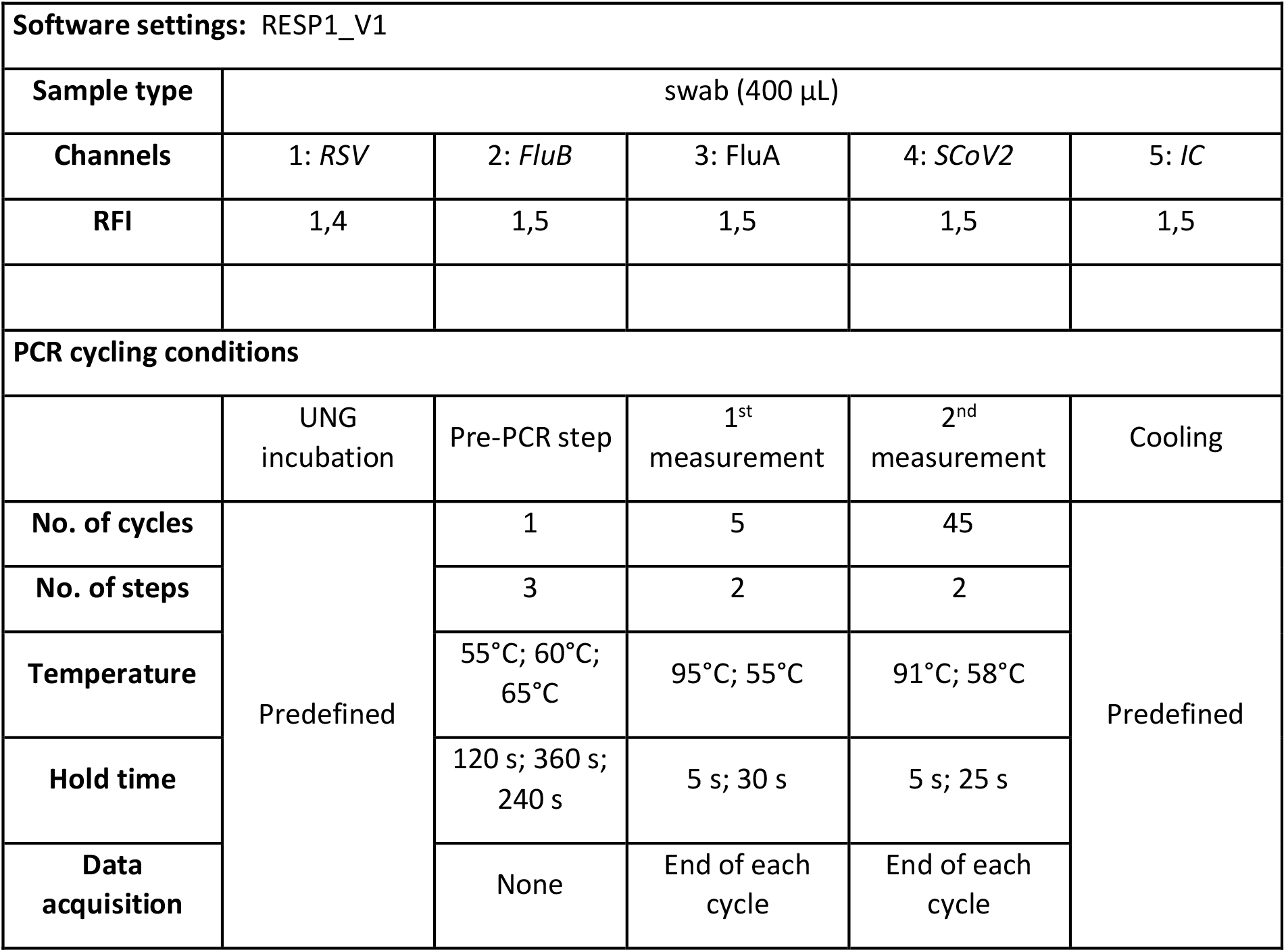
Software settings and run protocol: Cobas omni Utility Channel software settings and run protocol. IC: internal control, RFI: relative fluorescence increase.

| Software settings: RESP1_V1 |  |  |  |  |  |
| --- | --- | --- | --- | --- | --- |
| Sample type | swab (400 µL) |  |  |  |  |
| Channels | 1: RSV | 2: FluB | 3: FluA | 4: SCoV2 | 5: IC |
| RFI | 1,4 | 1,5 | 1,5 | 1,5 | 1,5 |
| PCR cycling conditions |  |  |  |  |  |
|  | UNG incubation | Pre-PCR step | 1<sup>st</sup> measurement | 2<sup>nd</sup> measurement | Cooling |
| No. of cycles | Predefined | 1 | 5 | 45 | Predefined |
| No. of steps |  | 3 | 2 | 2 |  |
| Temperature |  | 55°C; 60°C; 65°C | 95°C; 55°C | 91°C; 58°C |  |
| Hold time |  | 120 s; 360 s; 240 s | 5 s; 30 s | 5 s; 25 s |  |
| Data acquisition |  | None | End of each cycle | End of each cycle |  |

**Table 2.**
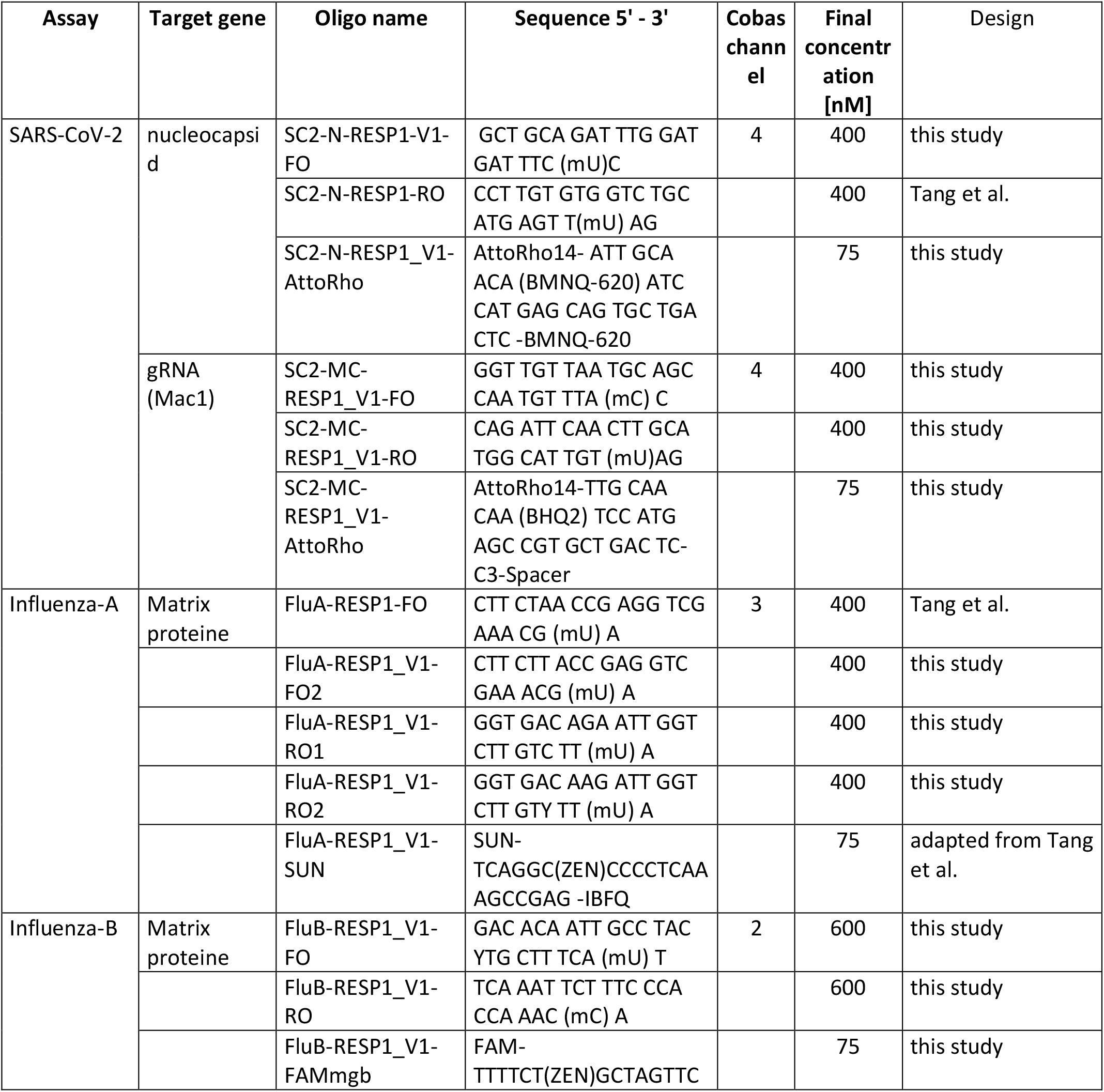
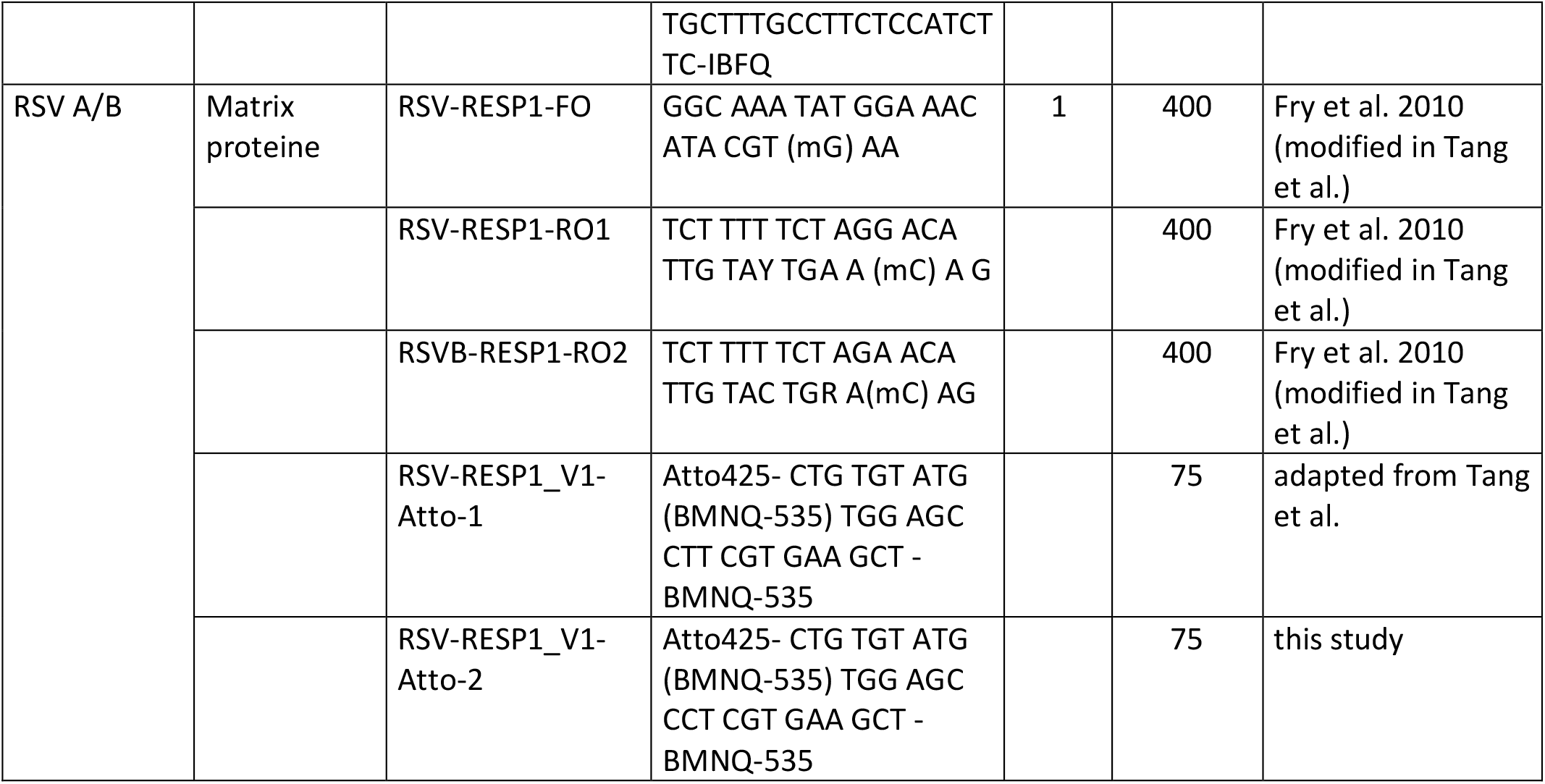
Details of assays and oligos for RESP1-V1. All oligos with modifications compared to RESP1 are highlighted in light green. Oligos that are sequence-identical but have a different fluorescent dye in RESP1_V1 are highlighted in light blue. Atto425, AttoRho14, FAM, SUN, YAK and Atto620 are different fluorescent dyes that allow detection of each target in one of the four *cobas5800/6800/8800 channels. mX: 2’O-methyl-RNA bases, ZEN/IBFQ: ZEN–Iowa Black fluorescence quencher, BHQ1: Black Hole Quencher 1, mgb: minor groove binder. Of note, the internal full-process control (added by default) uses Cobas channel 5 as described [7]*. Selected primers and probes were custom made by Ella Biotech GmbH (Fürstenfeldbruck, Germany), biomers.net (Ulm, Germany) and IDT (Coralville IA, USA).

### Analytical performance

Technical evaluation followed EU IVDR (2017/746). Quantitative standards were leftover diagnostic samples (FluA/B, RSV) quantified by digital-PCR as described [7], or WHO-standard (SARS-CoV-2). Serial two-fold dilutions (8 steps, 21 repeats) determined LoD (95% probit analysis). Linearity was assessed by ten-fold serial dilutions (5-6 steps, 5-10 per dilution) using Validation Manager software (Finbiosoft). Precision was determined in triplicates across three independent replicates. Cross-reactivity was analyzed using 36 clinical samples positive for viruses (n=19), bacteria (n=14) or fungi (n=2) (supplementary table 1). External quality assessment panels validated inclusivity (Instand, Germany).

### Clinical performance

RESP1_V1 was compared to the former assay (n=208 samples), then against cobas Respiratory flex test (Roche, Switzerland) using routine samples (n=167, winter 2024/2025 for FluA/B/RSV; until September 2025 for SARS-CoV-2). Both assays ran on cobas 8800. The comparator assay was used as instructed by the manufactorer.

## Results

### RESP_V1

The reevaluation led to substantial oligonucleotide modifications (Table 2). For SARS-CoV-2, we maintained the dual-target approach but replaced the spike gene due to high mutation rates. The macrodomain (Mac1) was identified as a promising target given its genetic stability [11]. Primers and probe oft he Mac1 assay show no mismatches with any variants (including XFG, NB1.8.1, LP.8.1.1) (Figure 1A) and presumably remain SARS-CoV-2 specific (>8 primer mismatches, 4 probe mismatches with SARS-CoV). The nucleoprotein was maintained with modified forward primer and probe. Adding Mac1 preserved robust amplification (Figure 1B-D).

**Figure 1:**
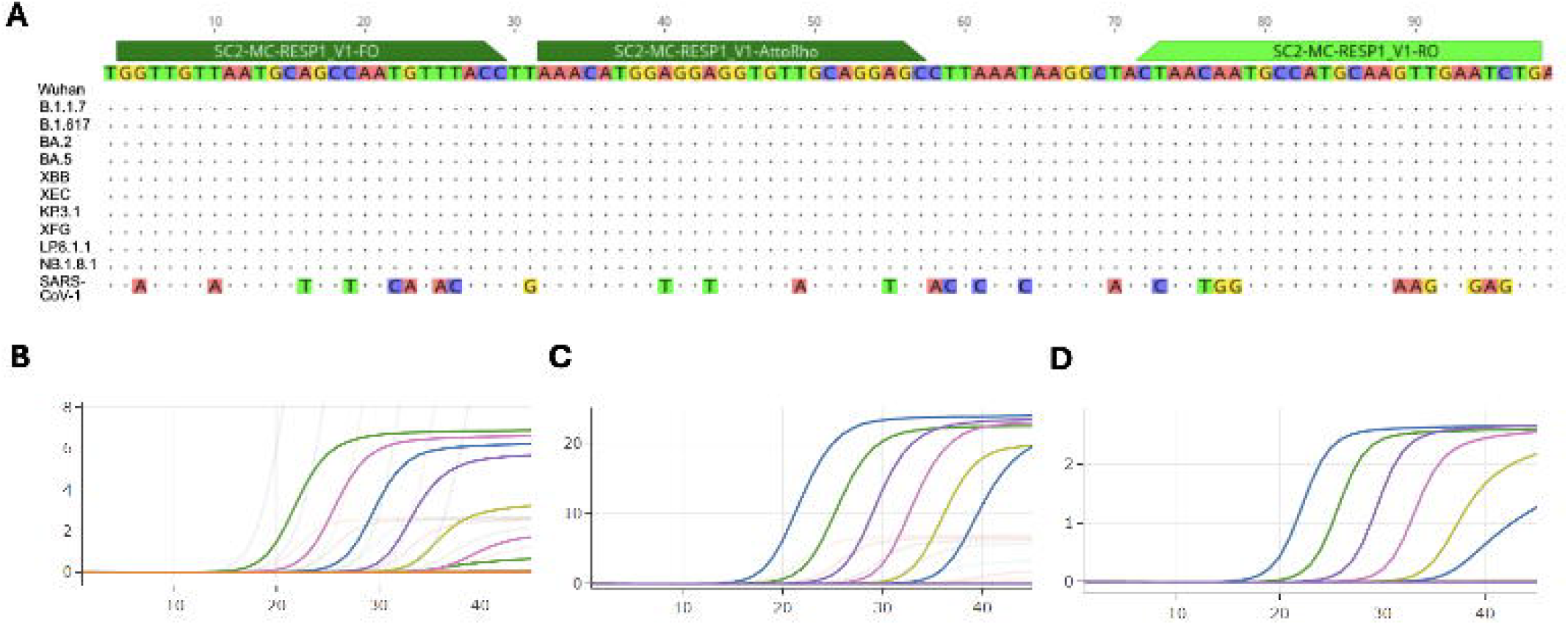
A) Representative alignment of the Mac1 region targeted by RESP1_V1 in indicated SARS-CoV-2 variants and SARS-CoV (NC_045512, OZ303875, PX393515, PV950741, PX461500, PQ016349, PX463174.1, PV950788, PX467198, PX467519, PX467497.1, NC_004718). B-D) Amplification curves of serial ten-fold dilutions of a representative clinical SARS CoV-2 positive sample tested in parallel with original LDT assay (B) and new version (RESP1_V1) with signals for the N (C) and Mac1 (D) target shown separately. Ct-values (x-axis) and relative fluorescence intensity (y-axis) are illustrated. Both the slightly modified SARS-CoV-2 N assay (C) and the new Mac1 assay (D) preserved amplification efficasy at lower concentrations.

For influenza A (matrix gene target), an additional forward primer and two new reverse primers were introduced for increased inclusivity. The FluA probe employs a different dye (SUN, 546-566 nm). The FluB assay was completely redesigned (matrix region target), showing significantly improved performance (Figure 2). An additional probe was added to the RSV assay (modified by Fry et al. [12]) to increase signal.

**Figure 2.**
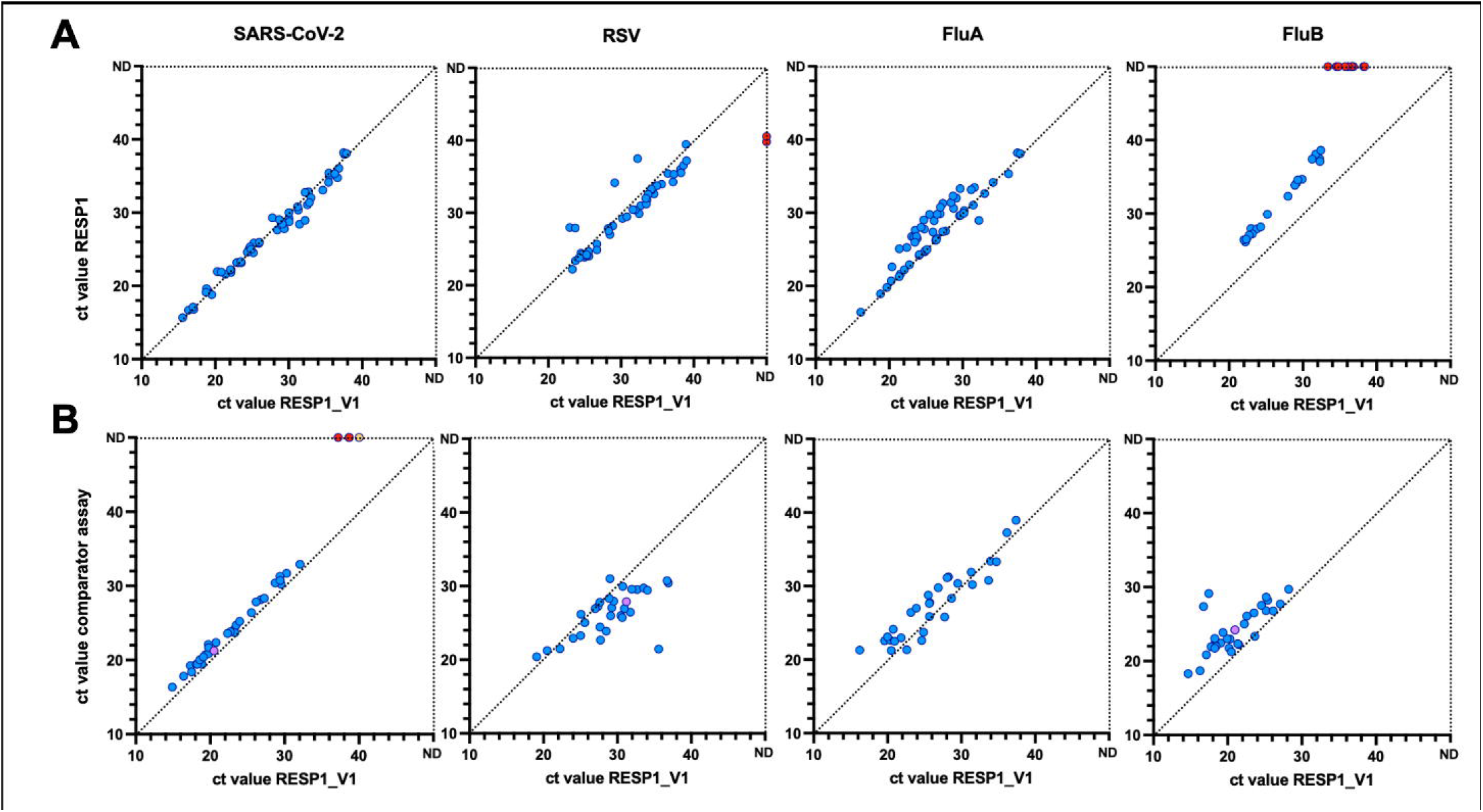
Clinical performance evaluation: A) Comparison of RESP1_V1 (x-axis) and RESP1 (y-axis), correlations of ct-values per assay are given. B) RESP1_V1 (x-axis) compared to the CE-IVD Respiratory Flex Test (y-axis). Samples that were negative in one of the two assays are shown as red dots, samples with dual detections (SARS-CoV-2, RSV, FluB) are shown in purple. Dashed diagonal line: Ideal linear correlation.

### Evaluation of the diagnostic performance

The analytical sensitivity analysis revealed RESP1_V1s LoDs to be within the range of RESP1 or lower, with SARS-CoV-2 70.9 (49.6-119) IU/ml, FluA H1N1 472 (314-913) cp/ml, FluA H3N2 112 (73.2—235) cp/ml, and RSV 1720 (1310-2740 cp/ml), repectively. RESP1_V1 showed excellent linearity for all four targets (r^2^: 0.989 – 0.997, linear ranges: 26.3-36.4/ 26.8-37.7/ 28.5-37.9/ 25.6-38.2 ct for SARS-CoV-2/FluA/FluB/RSV) (supplementary figure 1). Precision was high with (<0.58/<0.79/<0.75/<0.48 ct for SARS-CoV-2/FluA/FluB/RSV). Negative samples remained negative across all assessments. No false positives occured in the cross-reactivity panel (supplementary table 1) and EQA-panels were correctly detected (supplementary table 2).

#### Clinical evaluation

First, a set of n= 208 clinical samples was assessed in parallel by RESP1 and RESP1_V1 (n=171 positives, thereof n= 58/50/21/42 SARS-CoV-2/FluA/FluB/RSV positives). Good agreement was seen for SARS-CoV-2 and RSV. In RESP1_V1, detection of FluA/B was improved by mean 1.6/4.81 ct (Figure 2A).

Second, additional n=167 current clinical samples (thereof n= 117 positive (n=30 SARS-CoV-2, n=29 FluA, n=29 FluB, n=29 RSV), n=50 negative samples) were used for performance comparison to the CE-IVD assay on the cobas system. Overall, a high positive/negative agreement (94%/96% for SARS-CoV-2, 100%/100% for FluA/B, RSV) was observed. The lower SARS-CoV-2 agreement reflects n=4 samples with weak positive signals in RESP1_V1 (ct 37.19-40). Both tests correctly identified RSV in these samples (ct 22-34/21-29 in RESP1_V1/comparator). Retesting was impossible (samples depleted), thus false positives or true low-level detections cannot be discriminated, and contamination cannot be ruled out.

On a semiquantitative level, the SARS-CoV-2 assay shows comparable performance. For RSV, RESP1_V1 shows delayed detection versus the CE-IVD assay, whereas RESP1_V1 performed better for FluA and FluB (Figure 2B).

## Discussion

The adapted assay RESP1_V1 demonstrates robust analytical and clinical performance for simultaneous detection of SARS-CoV-2, influenza A/B, and RSV on a fully automated platform. By redesigning primer-probe sets we aimed to mitigate the risk of variant-driven performance loss while maintaining compatibility with automated high-throughput. The Mac1 domain of SARS-CoV-2 proved particularly valuable as a new diagnostic target. Its functional importance in viral replication and immune evasion imposes strong evolutionary constraints [9–11,13,14] reducing the likelihood of mutations that could compromise assay sensitivity. Our data confirm detection across contemporary SARS-CoV-2 lineages [15–19], validating this target selection strategy. A notable finding was the improved analytical sensitivity for recent influenza A and influenza B viruses compared to our previously published assay.

Clinical performance was challenged against a recently released CE-IVD assay (Respiratory Flex Test) on the same automated system. We demonstrated excellent correlation between the two tests. Notably, both SARS-CoV-2 assays showed a perfect linear correlation. Furthermore, the LDT demonstrated superior performance in the detection of influenza B.

In general, the focused syndromic testing approach provides practical advantages during respiratory season peaks when co-infections complicate differential diagnosis. Simultaneous detection streamlines laboratory workflows and accelerates clinical decision-making [20]. Integration into the fully automated platform preserves the operational advantages critical for routine diagnostics: high throughput, automated sample-to-result processing, and minimal hands-on time. The employment of LDTs with expertise in assay development facilitates swift response and provision of critical diagnostics [21,22]. fostering pandemic preparedness and ensuring autonomy from commercially oriented decisions.

Limitations include the need for ongoing genomic surveillance as viral populations continue to evolve. While in particular our SARS-CoV-2 assay design prioritizes functionally constrained regions, rare mutations may still emerge. Additionally, clinical validation was performed at a single site, and performance in diverse clinical contexts and geographical regions warrants further evaluation. Moreover, we can not rule out that a selection bias might have favored our LDT assay.

In conclusion, the updated assay balances analytical robustness, operational efficiency, and features an innovative target that might enhance resilience to SARS-CoV-2 evolution in the future. The approach demonstrates that periodic reassessment and strategic redesign of molecular diagnostics can maintain performance standards despite continuous viral evolution, supporting both routine clinical care and public health surveillance objectives.

## Author contributions

S.P. and M.L. conceptualized and supervised the study. D.N., K.G., H.T.T., L.S.P. and M.G. performed the experiments and data analysis. S.P. wrote and edited the manuscript. M.L., M.A. and S.P.discussed the data and corrected the manuscript. All authors have seen an approved the manuscript and agreed to its publication.

## Ethics and funding

This work was conducted in accordance with §12 of the Hamburg hospital law (§12 HmbKHG). The use of anonymized remnant diagnostic samples from patients was approved and informed consent was waived by the ethics committee of the Hamburg Medical Association (PV5626). The study was funded through the Network of University Medicine (NUM-SAR, funding code 01KX2524), and SP received funding by the Deutsche Forschungsgemeinschaft (DFG) (grant number 335447717; CRC1328, project A18). M.L and S.P. are associated to the DFG funded CRC1468.

## Data availability

The datasets generated and analysed during the current study are available from the corresponding authors on reasonable request.

