## supplement for "Refined fully automated RT-qPCR assay for simultaneous detection of SARS-CoV-2, Influenza A/B, and RSV with target optimization for improved variant resilience"

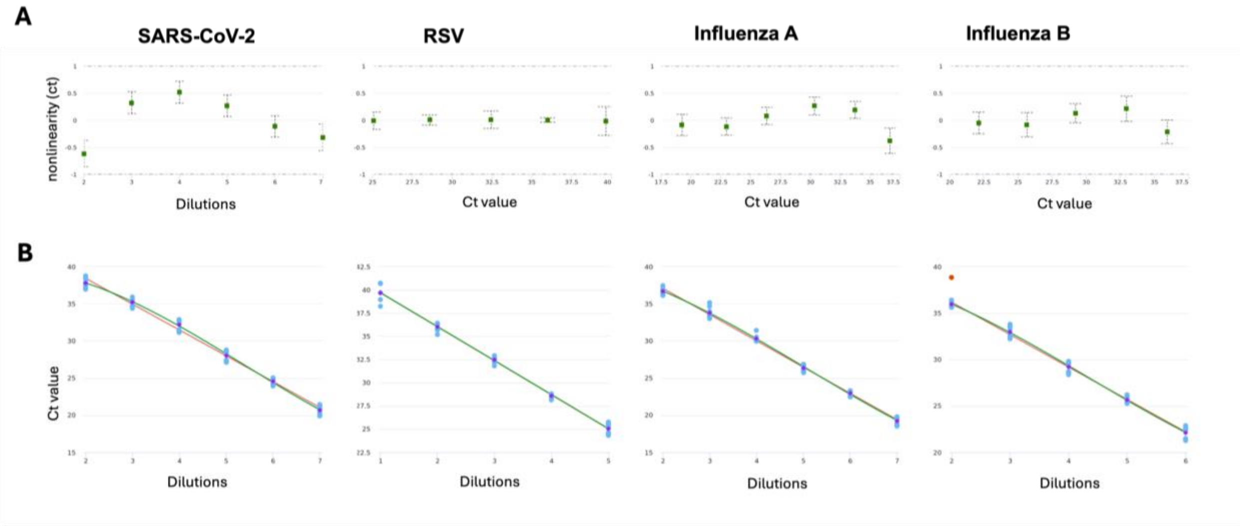


**Supplementary figure 1:** Analytical performance evaluation: Excellent linearity was observed for RESP1_V1. A) Nonlinearity is within the tolerable range (+/- 1 ct, y-axis) for all assays. The dashed lines indicate the 95% confidence interval. B) Linearity across the dilutions assessed. Blue dots correspond to the individual measured values, purple dots reflect the mean, red dots indicate outliers. Red line: linear fit; blue line: polynomial fit.

| | | RESP1_V1 | | | |
|---|---|---|---|---|---|
| sample number | Species detected in routine diagnostic | SARS-CoV-2 | Flu-A | FluB | RSV |
| 1 | Adenovirus | ND | ND | ND | ND |
| 2 | BK-Virus | ND | ND | ND | ND |
| 3 | BK-Virus | ND | ND | ND | ND |
| 4 | Bocavirus | ND | ND | ND | ND |
| 5 | CMV | ND | ND | ND | ND |
| 6 | EBV | ND | ND | ND | ND |
| 7 | EBV | ND | ND | ND | ND |
| 8 | HHV6 | ND | ND | ND | ND |
| 9 | HHV8 | ND | ND | ND | ND |
| 10 | hMPV | ND | ND | ND | ND |
| 11 | HPV | ND | ND | ND | ND |
| 12 | HSV-1 | ND | ND | ND | ND |
| 13 | HSV-2 | ND | ND | ND | ND |
| 14 | HSV-2 | ND | ND | ND | ND |
| 15 | human Coronavirus | ND | ND | ND | ND |
| 16 | JC-Virus | ND | ND | ND | ND |
| 17 | Parvovirus B19 | ND | ND | ND | ND |
| 18 | Rhino-Enterovirus | ND | ND | ND | ND |
| 19 | VZV | ND | ND | ND | ND |
| 20 | N. gonorrhoeae | ND | ND | ND | ND |
| 21 | Citrobacter koseri | ND | ND | ND | ND |
| 22 | Corynebacterium striatum | ND | ND | ND | ND |

| | | | | | |
|---|---|---|---|---|---|
| 23 | E. cloacae complex | ND | ND | ND | ND |
| 24 | E. faecium (VRE) | ND | ND | ND | ND |
| 25 | Enterococcus sp. | ND | ND | ND | ND |
| 26 | Haemophilus influenzae | ND | ND | ND | ND |
| 27 | K. oxytoca | ND | ND | ND | ND |
| 28 | Morganella morganii | ND | ND | ND | ND |
| 29 | Mycoplasma genitalium | ND | ND | ND | ND |
| 30 | Streptococcus agalactiae | ND | ND | ND | ND |
| 31 | Streptococcus pneumoniae | ND | ND | ND | ND |
| 32 | Streptococcus sanguinis grp | ND | ND | ND | ND |
| 33 | Ureaplasma parvum | ND | ND | ND | ND |
| 34 | Aspergillus sp. | ND | ND | ND | ND |
| 35 | P. jirovecii | ND | ND | ND | ND |

**Supplementary table 1.:** Cross reactivity assessment: Cross reactivity was tested on a set of clinical samples. The samples had previously tested positive for the specified pathogens as part of routine diagnostics. The new RESP1_V1 assay showed no signal in any of the samples. ND = not detected.

| Assay | strain/variant as given in EQA panel |
|---|---|
| SARS-CoV-2 | B.1, BetaCoV/Berlin/ChVir1670/2020_isolateBER |
| | B.1, BetaCoV/Munich/ChVir984/2020_IsolateBER |
| | B.1.351, BetaCoV/SouthAfrica/ChVir22131/2020 |
| | B.1.617.2 – hCoV-19/Germany/SHChVir25702_4/2021 |
| | BA.1, BetaCoV/Berlin/ChVir1670/2020 IsolatBER |
| | BA.1.1.7, BetaCoV/Passau/ChVir21652/2020 |
| | BA.2 – v8_26729_2_V.V.p7 |
| | JN.1 – v192_44042_V.V.p2 |
| | KP.3.1.1, V214_D24-06432_A.V.p2 |
| inluenza A | H1N1 (patient isolate 2024) |
| | H1N1 (patient isolate 2025) |
| | H1N1 pdm09 (A/Wisconsin/588/2019) |
| | H3N2 (A/Cambodia/e0826360/2020) |
| | H3N2 (patient isolate 2025) |
| | H5N1 (A/Whooper Swan/R65/2006) |
| | H5N1 clade 2.3.4.4b (A/wild goose/Germany-NW/AI00581/2024) |
| influenza B | B/Austria/1359417/2021 |
| | patient isolate 2025 |
| RSV | RSV A (patient isolate 2024) |
| | RSV A (patient isolate 2025) |
| | RSV B (patient isolate 2024) |
| | RSV B (patient isolate 2025) |

**Supplementary table 2:** Overview of the virus genomes contained in the EQA panels (starting in November 2024 through September 2025), all of which could be correctly detected using RESP1_V1.
